# Towards Automated Neonatal EEG Analysis: Multi-Center Validation of a Reliable Deep Learning Pipeline

**DOI:** 10.1101/2025.10.16.25338113

**Authors:** Tim Hermans, Anneleen Dereymaeker, Katrien Lemmens, Katrien Jansen, Fatima Usman, Shellie Robinson, Gunnar Naulaers, Maarten De Vos, Caroline Hartley

## Abstract

**Objective:** To evaluate the reliability and generalization of NeoNaid, a fully automated software tool for neonatal EEG analysis, based on functional brain age (FBA) estimation and sleep staging.

**Methods:** NeoNaid combines a multi-task deep learning model with proposed quality control routines detecting artefacts, out-of-distribution inputs, and uncertain predictions. Based on a raw EEG input, it outputs one global FBA estimate and a continuous 2-state hypnogram. We validated performance on an two independent hospital settings: an internal dataset (33 EEGs, 17 infants, median 900 minutes/recording) and an external dataset (38 EEGs, 24 infants, median 124 minutes/recording).

**Results:** Quality control rejected comparable number of segments in the internal and external datasets, reducing extreme errors in FBA estimation, and modestly improving sleep staging accuracy. Across the internal and external data, NeoNaid achieved median absolute FBA errors of 0.50 and 0.55 weeks and Cohen’s Kappa values of 0.89 and 0.87 for quiet sleep detection, respectively.

**Conclusions:** NeoNaid demonstrated improved reliability through integrated quality control and robust generalization across recording setups.

**Significance:** By focusing on validation and trustworthiness, this work takes an essential step toward clinical adoption of automated neonatal EEG analysis and supports its utility for both NICU practice and large-scale research.

## 1 Introduction

Neonatal electroencephalography (EEG) is the gold standard for assessing brain function in newborns and has proven to be a valuable monitoring tool in the neonatal intensive care unit (NICU) (Karamian & Wusthoff, 2021; McCoy & Hahn, 2013). With its high temporal resolution and multi-channel recordings, EEG provides rich information on brain maturation, sleep stages, and pathological activity such as seizures. However, neonatal EEG is challenging to interpret due to the complexity of the signal, and recordings often span many hours. Therefore, visual analysis requires expertise and is time-consuming. These factors limit the routine clinical use of EEG, despite its potential to provide valuable insights into neonatal brain health.

Over the past decade, a range of data-driven and artificial intelligence (AI) methods have been developed to assist with neonatal EEG interpretation. These include automated approaches for seizure detection (Ansari et al., 2019; Borovac et al., 2022; Raeisi et al., 2022; Temko et al., 2015), background grading (Dereymaeker et al., 2019; Moghadam et al., 2021; Raurale et al., 2021), sleep staging (Ansari et al., 2020; Koolen et al., 2017; Piryatinska et al., 2009), and functional brain age (FBA) estimation (Ansari et al., 2024; Pillay et al., 2020; Stevenson et al., 2020). These developments highlight the potential of artificial intelligence to scale EEG analysis beyond the limits of manual review.

Of the various AI approaches to neonatal EEG, sleep staging and FBA estimation are especially informative for assessing neurodevelopment, and are the focus of this study. Sleep organization is an important marker of neurological development (Dereymaeker et al., 2017; Shellhaas et al., 2017). Automated sleep staging can provide continuous, objective measurements that would otherwise be impractical for clinicians to obtain. Similarly, FBA estimation offers a quantitative measure of brain maturation by comparing EEG-derived estimates of age with the infant’s postmenstrual age (PMA). Deviations between the FBA and PMA can indicate atypical development and have prognostic value (Ansari et al., 2024; Stevenson et al., 2020). Together, these applications can support both clinical decision-making and long-term research on neonatal neurodevelopment.

Despite this potential, significant barriers remain to clinical adoption. Most published models are validated only on internal test data, raising concerns about their robustness to data from different hospitals, recording systems, or electrode montages. Long NICU recordings also unavoidably contain artefacts caused by movement, poor electrode contact, or physiological interference. Models trained primarily on clean data may fail when applied to such segments. For clinical usefulness, automated EEG tools must not only be accurate but also usable in practice and reliable across diverse datasets. Another important aspect for clinical deployment of automated EEG algorithms is their robustness to differences in recording setups. In neonatal care, EEG systems can vary in electrode montages, hardware characteristics, and acquisition protocols across centers. To be trusted in clinical workflows, algorithms must maintain performance despite these variations, including when applied to data from a hospital other than the one in which they were developed. Finally, to be adopted into clinical practice, tools must be easy to use with software that can aid interpretation.

To meet this need, we developed NeoNaid, a software tool that integrates a multi-task deep learning model for neonatal EEG analysis into a user-friendly graphical interface. Its design has been refined in discussion with clinicians to ensure interpretability and usability. This tool automatically processes long EEG recordings and provides robust, clinically relevant interpretations of the EEG, including sleep staging and FBA estimates. The underlying AI model builds on our previously published work and was trained on a large in-house dataset of neonatal EEG. Beyond this, NeoNaid implements quality control routines designed to improve the reliability and trustworthiness of the tool when used in clinical practice. These routines flag EEG segments likely to yield unreliable predictions by detecting artefacts, out-of-distribution inputs, or high model uncertainty.

In this paper, we focus on validating NeoNaid as a tool for neonatal EEG analysis. Unlike prior studies that primarily introduce new model architectures, our emphasis is on quality control and external validation. Specifically, we evaluate how NeoNaid performs on two independent datasets: an internal cohort from Leuven and an external cohort from Oxford. This cross-center validation is crucial for assessing generalizability and for building trust in real-world clinical use.

The aims of this study are twofold: (i) to assess the contribution of quality control to improving the reliability of automated FBA and sleep analysis, and (ii) to validate NeoNaid’s performance for FBA estimation and sleep staging across both internal and external datasets. Together, these analyses address the key requirements for clinical usefulness: trustworthiness and generalizability.

## 2 Materials and methods

### 2.1 Automated EEG analysis using NeoNaid

NeoNaid is an in-house developed software tool for automated neonatal EEG analysis. It integrates preprocessing, deep learning–based predictions, and quality control routines within a graphical user interface designed for clinical use (Figure 1). The input to NeoNaid is a raw multi-channel EEG recording, which may vary in montage depending on the acquisition system. NeoNaid can handle this variability, as the model processes each channel separately and uses an attention mechanism to combine channel-wise predictions into a global output.

**Figure 1.**
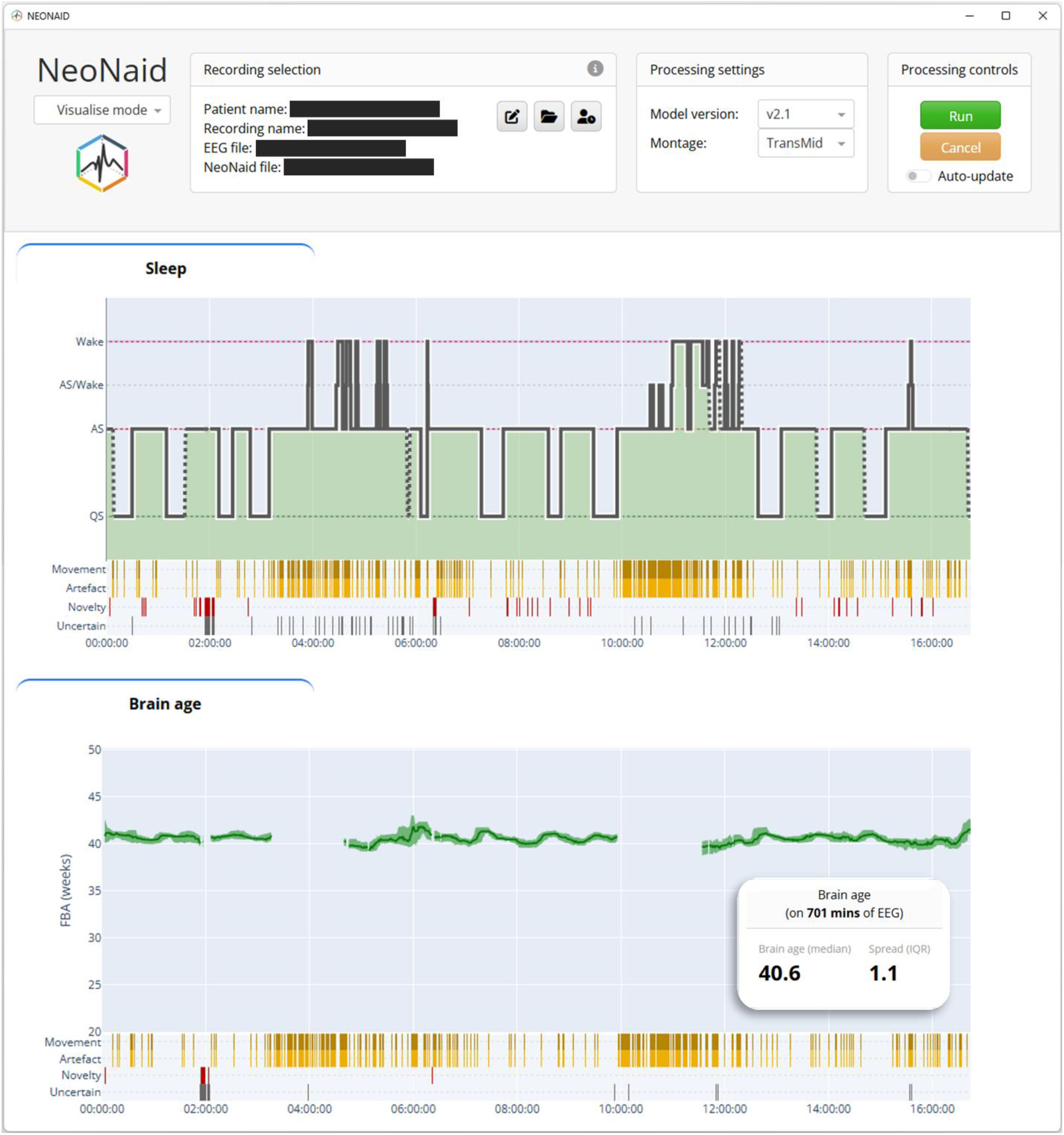
Screenshot of the NeoNaid GUI. The graphs show the predicted sleep hypnogram and the brain age as a function of time during a 16h45m recording. Besides these visual outputs, the GUI produces result files which were used to analyze the results for this paper.

Predictions are generated for non-overlapping 30-second segments. For each segment, NeoNaid produces three main outputs: (i) an artefact mask indicating which samples in the input are likely to be noise; (ii) a prediction of the sleep stage: quiet sleep (QS), active sleep (AS), or wake; and (iii) an estimate of functional brain age (FBA).

The core of the software is a multi-task deep learning model, developed by converging our previously published models (Ansari et al., 2019, 2020, 2024; Dereymaeker et al., 2019; Hermans et al., 2023) into one common methodology. Its architecture builds on our previously published convolutional neural network (Hermans et al., 2023) and consists of a shared encoder connected to multiple output heads, one for each task. Unlike prior single-task models, NeoNaid was trained in a multi-task setting, enabling simultaneous supervised learning from different neonatal EEG datasets labeled for different tasks. The training data included approximately 1326 hours of EEG from 124 recordings with age labels for FBA estimation, 565 hours from 132 recordings with sleep annotations, and 44 hours from 73 recordings with artefact annotations.

### 2.2 Dataset

We evaluated NeoNaid using two independent neonatal EEG datasets: one internal dataset collected at University Hospitals Leuven, Belgium (Dataset A) and one external dataset recorded at the John Radcliffe Hospital in Oxford, UK (Dataset B). These datasets differ in acquisition hardware, electrode configuration, and local recording protocols, allowing us to assess generalization of the algorithm to different recording conditions. The internal Dataset A originated from the same hospital as the data used for the development of the NeoNaid algorithms. However, all recordings Dataset A were independent of the development data and thus represent previously unseen cases.

#### 2.2.1 Dataset A (internal)

Dataset A consists of 33 EEG recordings from 17 neonates monitored at the NICU of University Hospitals Leuven, Belgium. The cohort includes both term and preterm infants with postmenstrual ages (PMA) at the time of recording ranging from 27.3 to 47 weeks (median 37.7). EEGs were acquired using the BrainRT EEG system (Onafhankelijke Software Groep (OSG), Kontich, Belgium) with a sampling rate of 250 or 256 Hz. Electrodes were positioned according to a modified 10–20 system, including the following channels: Fp1, Fp2, C3, C4, T3, T4, O1, and O2, with Cz as the reference electrode.

The EEG recordings had a median duration of 900 (IQR: 90 - 980) minutes and were collected as part of routine clinical care. All recordings came from neonates without major developmental abnormalities, allowing PMA at recording to serve as a reference measure for FBA. Two-class sleep annotations (QS versus AS or wake) were available for a subset of 28 recordings, scored by trained experts. All data were anonymized prior to analysis. The study was approved by the local ethics committee of University Hospitals Leuven, in accordance with the Declaration of Helsinki.

#### 2.2.2 Dataset B (external)

Dataset B comprises 38 EEG recordings from 24 neonates recorded at the Newborn Care Unit of the John Radcliffe Hospital, Oxford University Hospitals NHS Foundation Trust, Oxford, UK. The PMA at the time of recording ranged from 29.4 to 41.4 (median 34.5) weeks. Compared to Dataset A, Dataset B used a different EEG system and montage. EEGs were recorded using the SynAmps RT 64-channel headbox and amplifiers and CURRYscan7 neuroimaging suite (Compumedics Neuroscan) with a sampling rate of 2000 Hz. The electrode configuration consisted of FCz, C3, C4, Cz, CPz, T3, T4, and Oz, referenced to Fz.

Recordings had a median duration of 124 (IQR: 91 – 143) minutes. Similar to dataset A, all recordings were from patients without any neurological abnormalities (infants were excluded from studies if they had a grade III or IV intraventricular haemorrhage, hypoxic ischaemic encephalopathy, or major congenital malformations), making the PMA at time of recording a suitable reference for functional brain age. Of the 38 EEGs, 18 were randomly selected for sleep staging, ensuring good coverage across PMA and recording durations, and were subsequently labeled for sleep by a trained expert in Leuven.

The EEG recordings were collected for research purposes, as part of an independent study (Usman et al., 2025). Eligible families were given verbal and written information about the study, and written parental consent was signed before inclusion in the study. This dataset was fully anonymized and provided under a data sharing agreement between the University of Oxford and KU Leuven. Ethical approval was obtained through the relevant UK regulatory bodies (National Research Ethics Service reference: 12/SC/0447).

#### 2.2.3 Pre-processing

To standardize inputs across datasets, all recordings were re-referenced to a bipolar montage (C3–C4, C3–Cz, C3–T4, C4–Cz, C4–T3), bandpass-filtered (0.25–30 Hz), and downsampled to 64 Hz. Signals were segmented into 30-second non-overlapping epochs. Within each recording, channel amplitudes were normalized by the median standard deviation across all segments. These pre-processing steps ensured consistency across acquisition systems and were automatically executed by the NeoNaid software.

### 2.3 Quality control

Typically, the majority of training data is from a clean and labeled dataset, thus making existing trained models unreliable for artefact-containing data segments. Additionally, deep learning models perform well on data that is similar to the training data, but their performance becomes unreliable when applied to data that has significantly different characteristics compared to the training data (out-of-distribution data).

A central feature of NeoNaid is its quality control algorithm, which evaluates the reliability of each 30-second EEG segment before downstream interpretation. This process involves evaluating three independent reliability criteria: artefact content, novelty detection (to detect out-of-distribution inputs), and (un)certainty level.

#### 2.3.1 Artefacts

Each segment is assigned an artefact score based on the percentage of samples identified as noise by the dedicated detection head of the model. Segments with more than 50% artefact content are flagged as unreliable, therefore preventing the model from producing predictions on segments where brain activity is largely obscured. Segments that do not pass the artefact check are flagged as unreliable and either excluded (for FBA) or explicitly marked (for sleep staging).

#### 2.3.2 Novelties

Out-of-distribution inputs (novelties) are automatically identified by applying a novelty detection model to each channel in a segment. The novelty detection model in NeoNaid is an isolation forest that uses a set of nine spectral features as input (Piryatinska et al., 2009), predicting for each channel in every segment whether it is an inlier or a novelty (with respect to the NeoNaid training data). A separate novelty detection model was fitted per task, using the data that was used for training that part of the multi-task model. If more than half of the channels in a segment were labeled as novelties, the entire segment was flagged. Conversely, unflagged segments in any of the channels labeled as novelties were excluded from the model’s channel aggregation, reducing their impact on the global predictions.

#### 2.3.3 Uncertainty

The model assigns attention weights to each channel when combining channel-wise predictions. Very low weights across all channels indicate uncertainty. Segments in which the maximum attention weight was below a predefined threshold were therefore flagged as unreliable. For the sleep staging outputs, segments with QS probabilities near 0.5 were also marked as uncertain.

Together, these three criteria provide a conservative safeguard against unreliable predictions. NeoNaid then aggregates the segment-wise outputs and quality flags into clinically interpretable results. For FBA, a single robust estimate is obtained as the median across reliable segments, while for sleep staging, a continuous hypnogram is constructed by smoothing probabilities and interpolating over short, unreliable intervals using simple heuristic rules.

### 2.4 Performance metrics

#### 2.4.1 Functional brain age

For each recording, the global FBA estimate was defined as the median of all segment-wise predictions that passed quality control. The performance was quantified in terms of the absolute error, which was defined as the absolute difference between the global FBA estimate and the infant’s PMA at the time of recording. Lower errors indicate better performance. The interquartile range (IQR) of the retained segment-wise estimates was reported as a measure of prediction confidence, with wider IQRs indicating a lower prediction certainty.

#### 2.4.2 Sleep staging

For sleep analysis, we evaluated NeoNaid’s ability to detect QS. To this end, the model’s AS and wake predictions were combined into a single category, representing the non-quiet sleep class. Predictions on segments flagged as unreliable were excluded. The performance was measured in terms of Cohen’s kappa score, where a higher score indicates better agreement between predicted sleep stages and expert annotations.

### 2.5 Analysis

Our analyses were designed to evaluate both the impact of quality control and the generalizability of NeoNaid across datasets. We applied the full processing pipeline to both datasets: Dataset A (internal) and Dataset B (external). We compared two approaches: a naïve approach that included all segment-wise predictions, and a robust approach that excluded segments flagged by quality control.

#### 2.5.1 Effect of quality control

To check for differences in data quality and characteristics between the two datasets, we first quantified the occurrence of quality control flags within each dataset. More specifically, we computed the proportion of EEG segments flagged for artefacts and novelty detection; these were independently assessed for the FBA and sleep tasks.

For FBA estimation, performance was evaluated as a function of EEG recording duration, as the effect of the robust method is most clear for EEGs of shorter durations. To simulate varying EEG durations, we extracted sub-epochs ranging from 30 seconds to 1 hour from each recording. For each duration, 1000 sub-epochs were randomly selected per recording. The median FBA and corresponding performance metrics were calculated for each duration, allowing assessment of how recording length and the inclusion of quality control routines affect prediction errors.

For sleep staging, we computed Cohen’s kappa score for quiet sleep detection on the full recordings using both the naive and robust approaches and compared the performance.

#### 2.5.2 Cross-center validation

Finally, we validated the robust methodology (i.e., including quality control) on the complete EEG recordings. Per-recording results were analyzed and visualized in two ways. Firstly, FBA and sleep performance metrics are computed separately for each channel, using the per-channel predictions. A global result was generated using the weighted average of the per-channel predictions. Secondly, we showed the performance as a function of PMA to investigate whether prediction accuracy is systematically affected by the age of the neonates.

## 3 Results

### 3.1.1 NeoNaid quality control reduces errors

We investigated how the quality control routine affects the automated analysis. For FBA, the median rejection rate was 21.5% in the internal dataset and 16.1% in the external dataset (Figure 2). These were lower for sleep staging, with medians of 2.0% (internal) and 2.4% (external), mainly due to the heuristic postprocessing, which interpolates short unreliable intervals, and retains segments with high-amplitude movement artefacts when they occur during predicted wake cycle.

**Figure 2.**
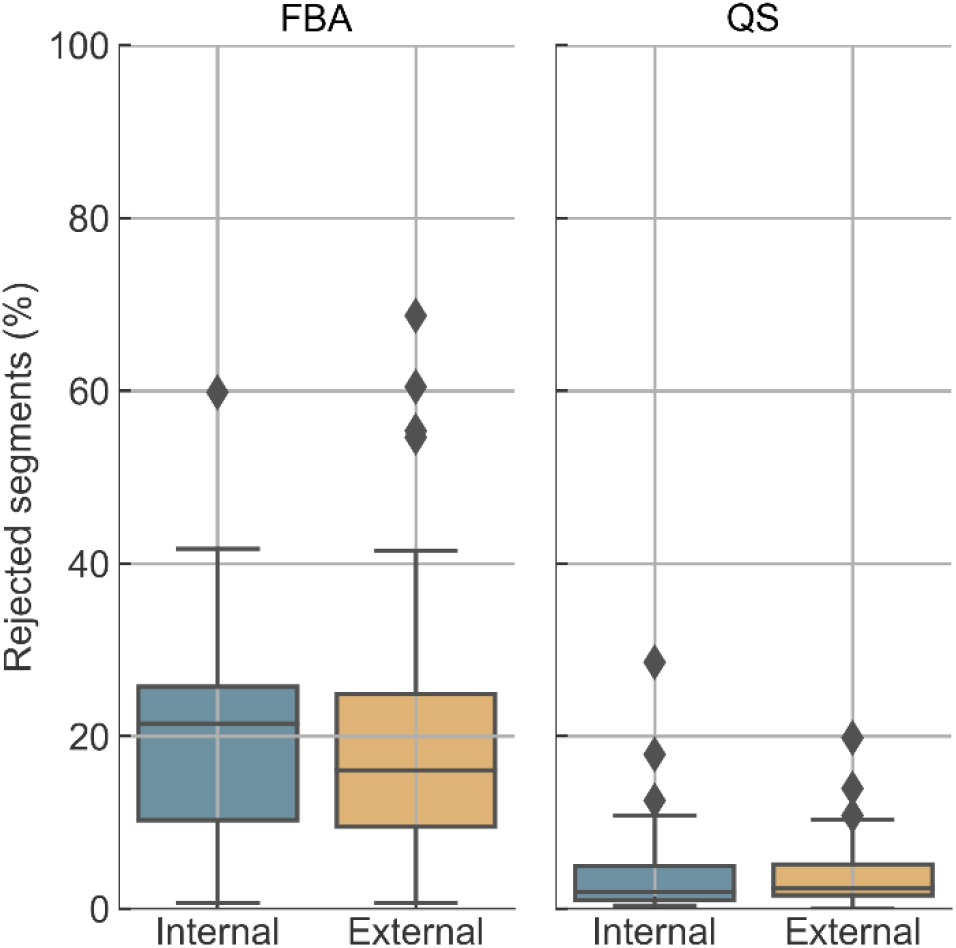
Rejection rates due to quality control in the two datasets. Rejected %: the total percentage of segments that were rejected in each recording.

Analyzing the rejection rates according to the three quality control criteria (which are not mutually exclusive), segments were most frequently flagged as artefacts. In the FBA data, 22.9% of segments in the internal dataset and 19.6% in the external dataset were marked as artefactual; similar rates were observed for sleep data (22.3% and 19.3%, respectively). Novelty detection contributed less, flagging 1.1% and 2.4% of FBA segments, and 4.1% and 3.4% of sleep segments in the internal and external data, respectively. The FBA segments uncertainty was 3.3% (4.6% sleep segments: internal) and 7.1% (2.2% of sleep segments: external). For sleep staging, these values are observed prior to heuristic postprocessing, thus accounting for the generally lower rejection percentages than the proportion of initially flagged segments. Overall, data quality was comparable between centers, and the external data did not appear out-of-distribution despite differences in acquisition systems and protocols.

Next, we investigated the effect of the quality control on FBA performance in both datasets. Overall, the robust method (which applies segment rejection) and the naive method (which does not) yielded similar median FBA error values (Figure 3). This is expected, as the use of the median as an aggregation metric reduces the influence of outlier segments. Nevertheless, the robust method consistently showed a lower likelihood of producing extreme outliers, particularly in shorter recordings, demonstrating its value in minimizing risk. The median IQR of the FBA estimate initially increased with data length and then plateaued after approximately 20 minutes of usable data. This suggests that IQR values in recordings with less than 20 minutes of non-rejected EEG should be interpreted with care, as limited data availability may underestimate the IQR of the actual underlying distribution.

**Figure 3.**
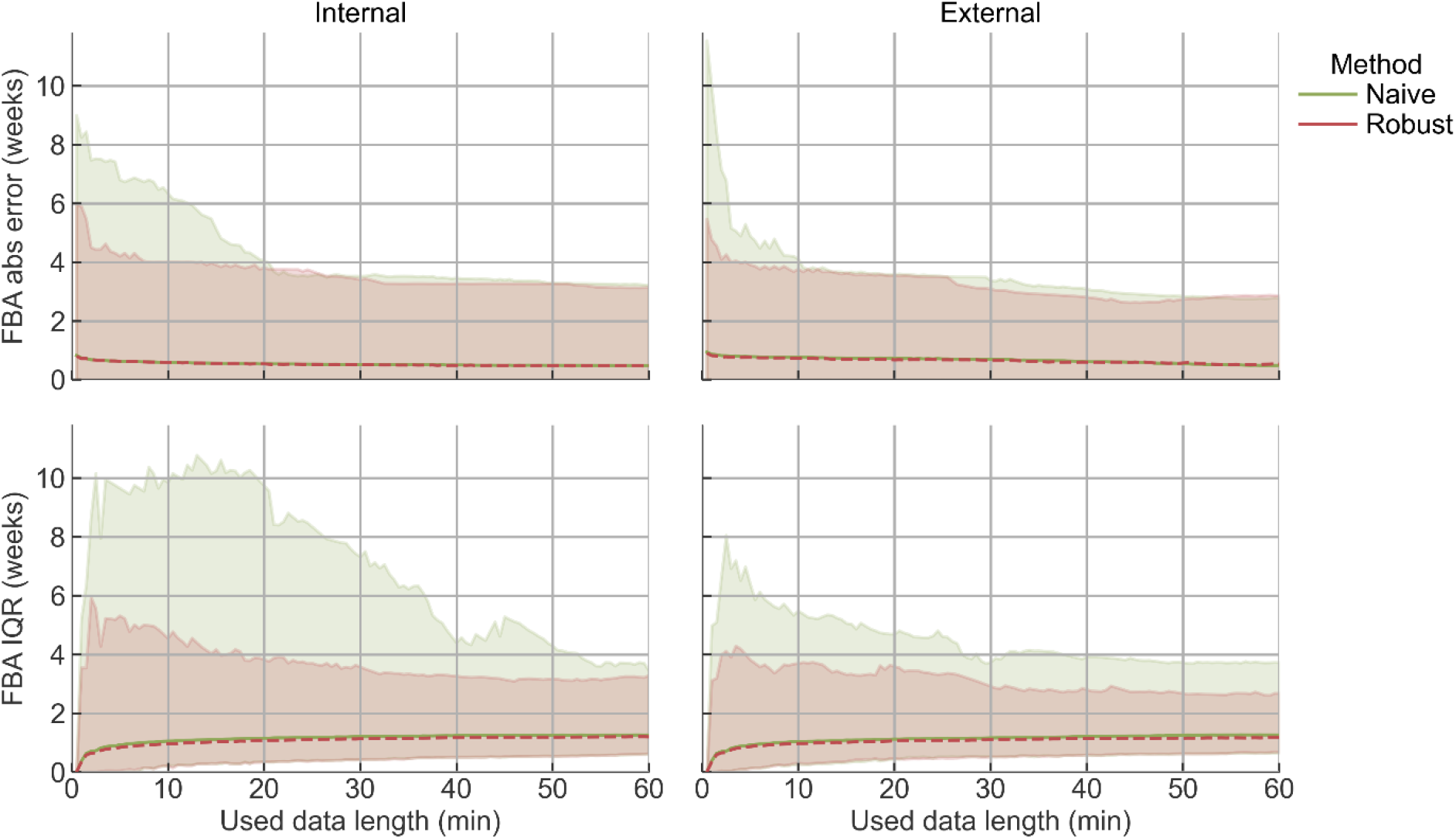
FBA performance for different simulated data lengths (1000 repetitions per window, per recording). The x-axis represents the duration of data used to compute the estimates (excluding the rejected segments in the robust case), of which **median + min-max** is shown. Robust method includes quality control and subsequent segment rejection, naïve method does not. Left: internal Dataset A, right: external Dataset B.

QS detection performance using the naive and robust methods (Figure 4) showed modest performance improvement following quality control on the already relatively cleaned and labeled sleep data. Unusable segments were not annotated by the experts and, therefore, were not included in the performance evaluation. While the benefit of quality control is less pronounced in this evaluation setup, its primary value lies in preventing unreliable predictions when poor-quality data are encountered in practice.

**Figure 4.**
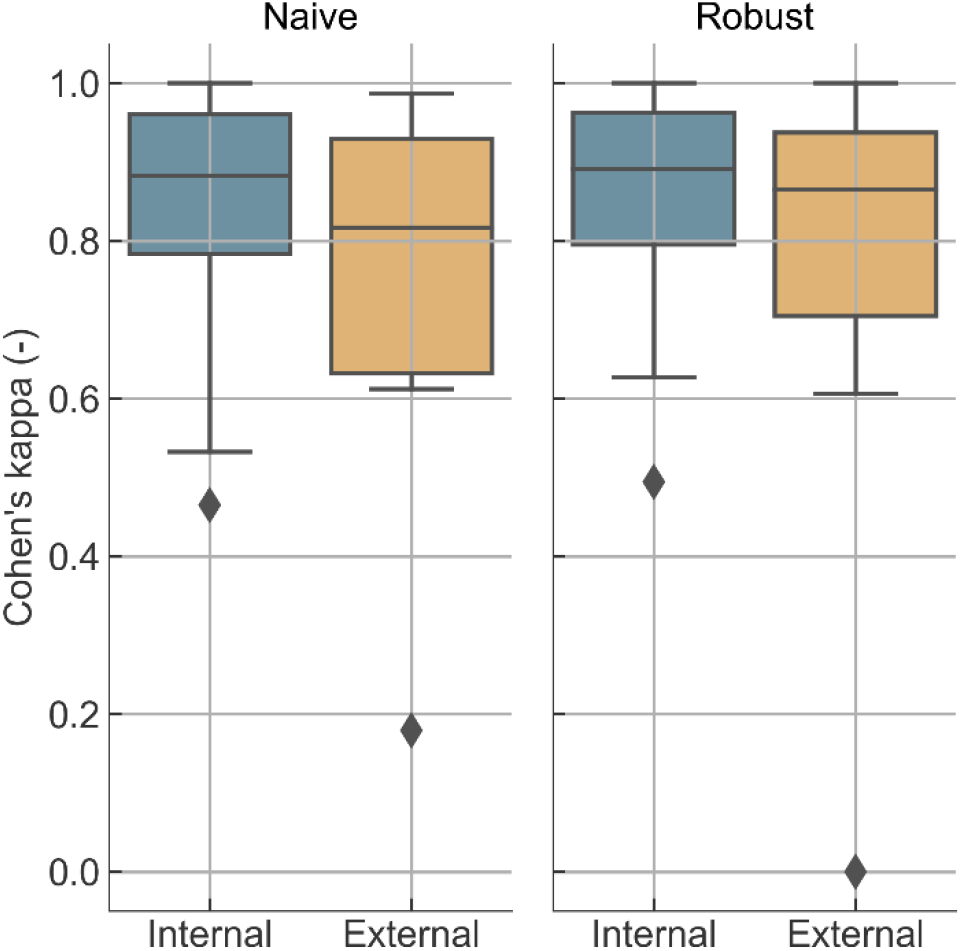
Sleep performance with (robust) and without (naive) artefact rejection and heuristic postprocessing.

### 3.1.2 Cross-center validation of NeoNaid

We investigated the results of the robust method (with quality control) of both datasets to validate the cross-center generalization of NeoNaid. Using the robust methodology on full recordings, the mean absolute FBA error across the datasets is 0.60 (median:0.50, IQR: 0.21-0.78) weeks for Dataset A, and 0.69 (median: 0.55, IQR: 0.23-1.02) weeks for Dataset B. The percentage of recordings within 1 week error is 78% (100% within 2 weeks) and 74% (93% within 2 weeks) for Dataset A and Dataset B, respectively. Moreover, the true PMA for Dataset A fell within the IQR of the per-segment predictions in 70% of recordings, compared to 58% for Dataset B.

For QS detection, performance was high in both datasets. In Dataset A, per-recording kappa scores averaged 0.86 (median: 0.89, IQR: 0.80-0.96), while in Dataset B they averaged 0.79 (median: 0.87, IQR: 0.70-0.94). Comparable results were obtained when restricting the analysis to single channels, particularly for bipolar derivations around C3, Cz, and C4 (Figure 5). When pooling all recordings, the overall kappa was 0.874 (95% CI: 0.865–0.884) in Dataset A and 0.831 (95% CI: 0.811–0.851) in Dataset B.

**Figure 5.**
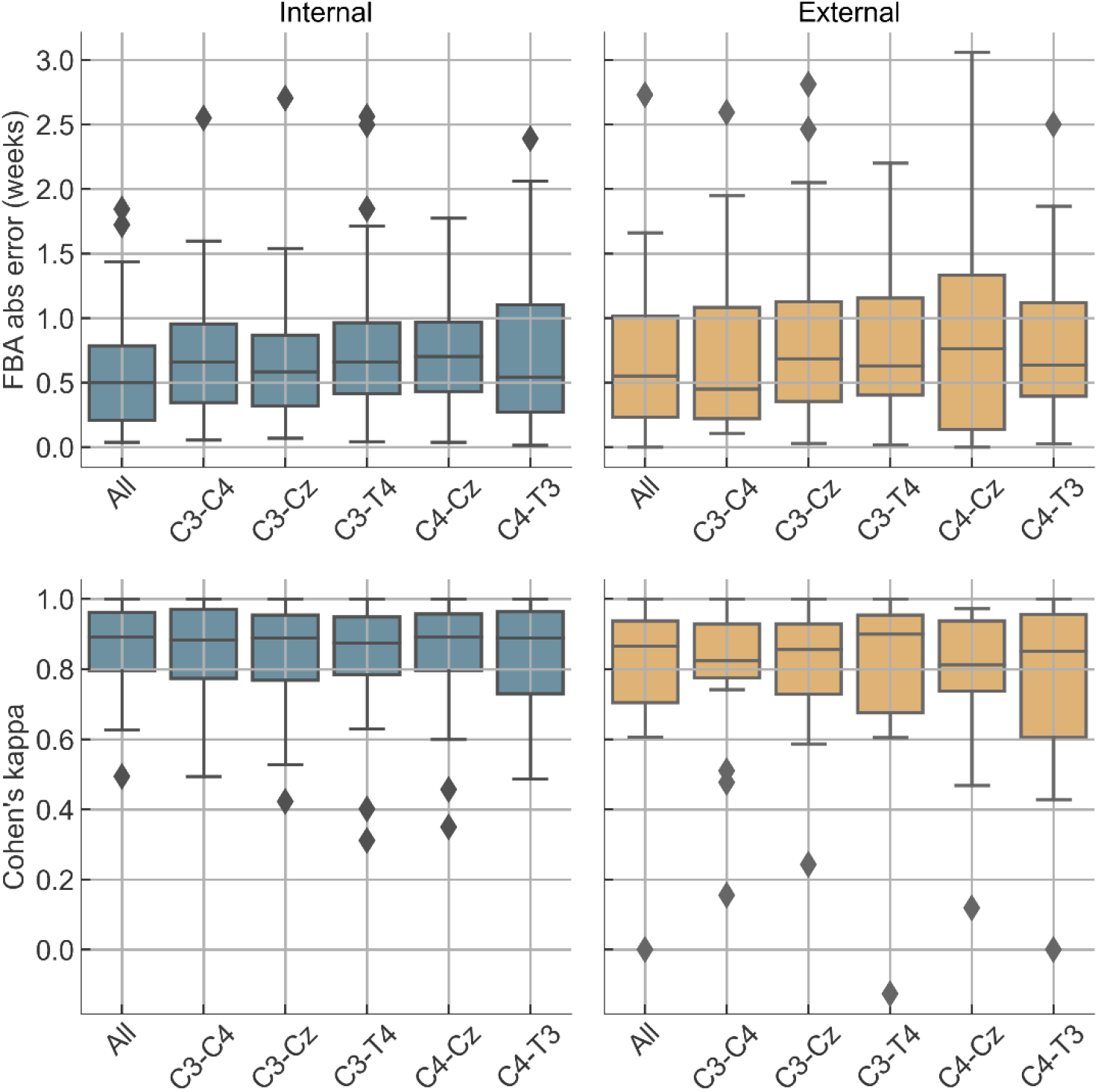
Per-recording performance per channel. “All” refers to attention-weighted average of single-channel predictions. Top: brain age error; bottom: quiet sleep detection score. Left: internal Dataset A, right: external Dataset B

The performance of FBA and quiet sleep detection was relatively consistent across channels (Figure 5), suggesting that the software can deliver reliable results even with limited or single-channel input. However, while combining all channels did not always outperform the best individual channel in terms of median performance, it generally helped reduce errors in outlier recordings, offering improved robustness.

Finally, we assessed the model performance compared with the age of the infants. For the FBA model, there is no significant linear correlation between the FBA error and PMA (Figure 6), indicating that the model performed equally at all ages. In contrast, for sleep staging, there was a trend that QS performance improves with increasing age (Figure 6). In one case in Dataset B, the model failed to detect any quiet sleep, whereas the expert annotation indicated a 15-minute QS bout, resulting in a Kappa score of zero. The 15-minute QS epoch included multiple high-amplitude artefacts, which resulted in the model misclassifying it as wake.

**Figure 6.**
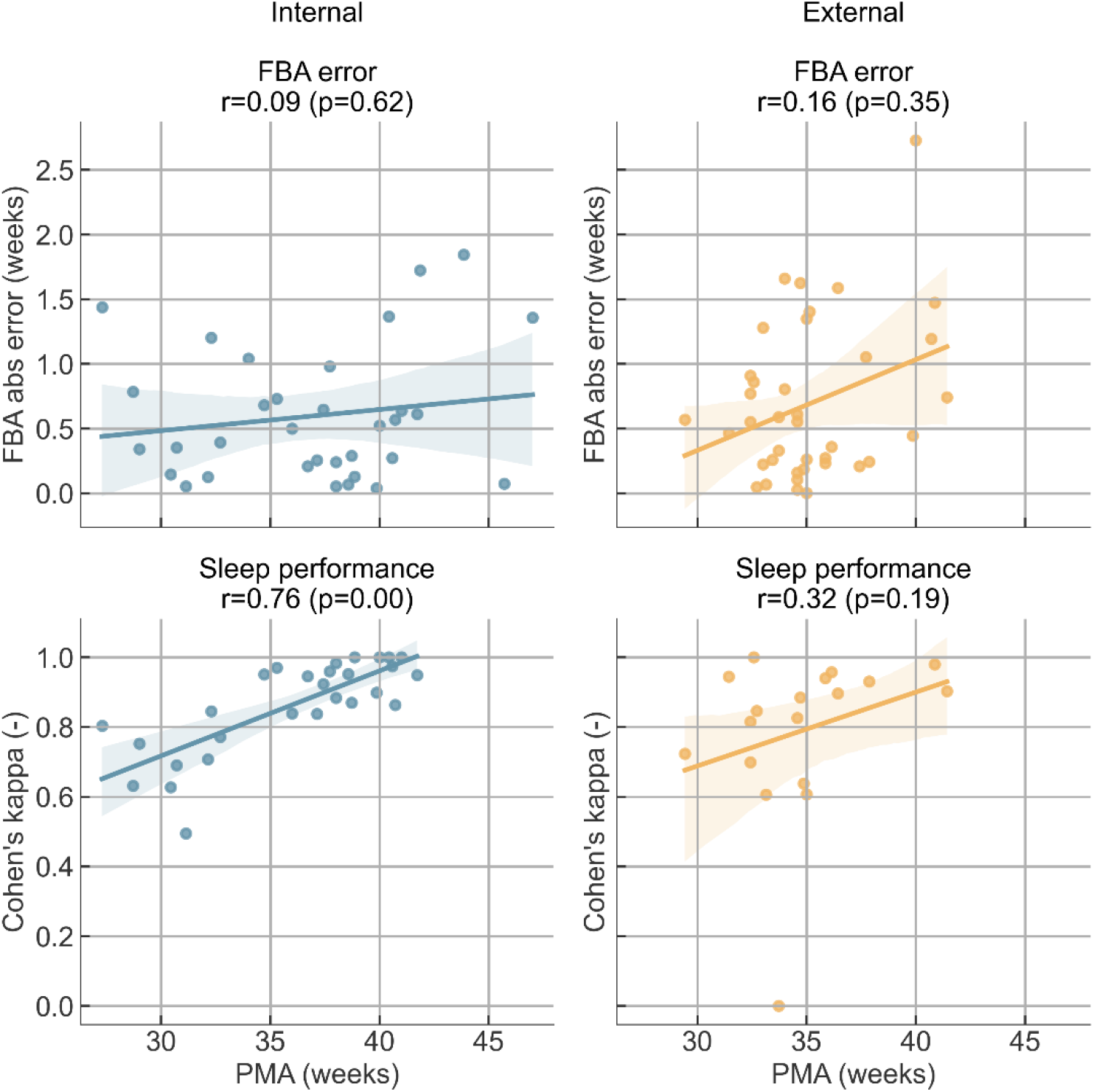
The relation between the performance and PMA. Left: internal Dataset A, right: external Dataset B.

## 4 Discussion

The aim of this study was twofold: first, to assess the contribution of quality control routines in improving the reliability of automated neonatal EEG analysis, and second, to validate the performance of NeoNaid across independent datasets from two hospital settings. We found that quality control reduced extreme errors and provided transparent confidence measures, particularly for functional brain age estimation in shorter or noisier recordings. Across both centers, NeoNaid demonstrated robust and generalizable performance for functional brain age and sleep staging, despite differences in recording hardware, montages, and protocols. This is an important step toward the broader clinical adoption of AI tools for neonatal EEG, where differences in acquisition setups are common and difficult to standardize.

NeoNaid’s integrated quality control framework assesses the reliability of each EEG segment through artefact detection, novelty detection, and attention-based certainty scoring. Our results show that this quality control framework improves the reliability of automated neonatal EEG analysis by reducing the likelihood of extreme errors while preserving valid information. While average performance metrics changed only modestly, the safeguards proved valuable in minimizing risk, particularly in shorter or lower-quality recordings (Figure 3 and Figure 4). Therefore, these routines increase the clinical trust in the outputs generated by NeoNaid.

NeoNaid maintained robust performance when validated on an independent, external dataset, despite differences in acquisition hardware, montages, and protocols. Additionally, the external data were not identified as out-of-distribution by the quality control routines. This cross-center validation highlights the generalizability of the tool and addresses a key barrier to clinical adoption of automated EEG methods. Even with single-channel inputs, NeoNaid maintained strong performance, producing comparable results compared to aggregating a 5-channel setup (Figure 5). By means of an attention-based aggregation across channels, the model puts more weight on channels it finds most reliable. Therefore, using more than one channel improves robustness more than accuracy, by not relying on data (quality) in a single channel.

Performance did not vary significantly with PMA for FBA, although there was a tendency toward slightly larger errors in recordings below 30 or above 40 weeks PMA. This pattern likely reflects the limited amount of training and validation data available in these extreme age ranges rather than a systematic bias. In contrast, sleep staging performance clearly improved with age. As shown in Figure 6, older neonates achieved higher kappa scores for quiet sleep detection, whereas younger infants showed slightly lower performance. This is likely due to the less distinct differentiation between active and quiet sleep at earlier developmental stages.

Several prior studies have developed deep learning models for brain age estimation (Ansari et al., 2024; Pillay et al., 2020; Stevenson et al., 2020) and sleep staging. While direct comparison of performance metrics across studies should be done cautiously due to differences in test datasets, our findings show that the FBA performance of NeoNaid (mean absolute errors of 0.60 and 0.69 on internal and external datasets, respectively) is comparable, if not superior, to these earlier models, which typically report mean absolute errors between 0.7 and 1.0 weeks. Similarly, NeoNaid’s performance in QS detection aligns with prior models that have reported kappa values up to 0.77.

NeoNaid offers value for both clinical and research applications. In clinical practice, automatic sleep staging and FBA estimation can aid in monitoring brain development, particularly in preterm infants. The IQR accompanying the FBA estimate provides a practical measure of confidence, helping clinicians interpret results more effectively. The built-in quality control indicators can alert users to unreliable segments, reducing the risk of misinterpretation due to artefacts or signal degradation. For researchers, NeoNaid provides a scalable solution for annotating large EEG datasets in a standardized way. It is especially useful in studies investigating neurodevelopmental trajectories, sleep–wake organization, and responses to therapeutic interventions.

This study has several limitations. The number of recordings in the external dataset was relatively small, limiting the statistical power of generalization claims. Sleep annotations were made by different raters at each site, without a formal assessment of inter-rater reliability, which could introduce bias. Moreover, the external recordings were shorter in duration and lacked accompanying physiological or video data, which made them more challenging to annotate. Finally, external validation was limited to a single center. Future work will focus on expanding collaborations to include additional external datasets and broader populations to further validate and refine the NeoNaid platform.

## 5 Conclusion

We have shown that NeoNaid is a robust tool for automated neonatal EEG analysis, achieving good performance and generalizability across datasets with different recording setups. Its integrated quality control routines reduce extreme errors and improve trustworthiness, addressing a critical requirement for clinical adoption. Together, these findings demonstrate that quality control and cross-center validation are essential for clinical reliability, and they support the potential of NeoNaid for both NICU practice and large-scale research on neonatal brain monitoring.

## Data Availability

All data produced in the present study are available upon reasonable request to the authors.

## Author contributions

CRediT roles: TH: Conceptualization, Methodology, Software, Validation, Writing - Original Draft. AD: Conceptualization, Methodology, Investigation, Writing - Review & Editing. KJ and GN: Conceptualization, Investigation. KL, FU and SR: Investigation. MDV and CH: Conceptualization, Methodology, Resources, Writing - Review & Editing, Supervision.

## Declaration of competing interest

The authors declare that they have no known competing financial interests or personal relationships that could have appeared to influence the work reported in this paper.

### Acknowledgements

This research received funding from i) the European Union as part of the Cost action “Maximising impact of multidisciplinary research in early diagnosis of neonatal brain injury” (AI-4-NICU) CA20124; ii) Industrieel Onderzoeksfonds KU Leuven (IOF): C3 “NEONAID - Neonatal Online Neuro-Analysis for Interpretation and Decision-support - Bringing automated neonatal monitoring to the bedside”, C3/24/023; iii) the Horizon programme: HORIZON-HLTH-2022-IND-13: “Privacy compliant health data as a service for AI development (PHASE IV AI)”, funded by the European Union, under Grant Agreement #101095384; iv) the Flemish Government (AI Research Program), MDV and TH are affiliated to Leuven.AI - KU Leuven institute for AI, B-3000, Leuven, Belgium; v) CH and SR were funded by the Wellcome Trust/Royal Society through a Sir Henry Dale Fellowship awarded to CH (grant reference: 213486/Z/18/Z). FU was funded by the Commonwealth Scholarship Commission.

## Declaration of generative AI and AI-assisted technologies in the writing process

During the preparation of this work, the authors used ChatGPT in order to improve readability and language in portions of the manuscript. After using this tool/service, the authors reviewed and edited the content as needed and take full responsibility for the content of the publication.

